# A Fundamental Distinction in Early Neural Processing of Implicit Social Interpretation in Schizophrenia and Bipolar Disorder

**DOI:** 10.1101/2021.03.08.21253057

**Authors:** Nuno Gonçalo Gomes Fernandes Madeira, Ricardo Filipe Alves Martins, João Valente Duarte, Gabriel Nascimento Ferreira Costa, António João Ferreira Macedo Santos, Miguel Sá Sousa Castelo-Branco

## Abstract

**Background:** Social cognition impairment is a key phenomenon in serious mental disorders such as schizophrenia (SCZ) and bipolar disorder (BPD). Although genetic and neurobiological studies have suggested common neural correlates, here we hypothesized that a fundamental dissociation of social processing occurs at an early level in these conditions.

**Methods:** Based on the hypothesis that key structures in the social brain, namely the temporoparietal junction, should present distinctive features in SCZ and BPD during low-level social judgment, we conducted a case-control study in SCZ (n=20) and BPD (n=20) patients and controls (n=20), using task-based fMRI during a Theory-of-Mind (ToM) visual paradigm leading to interpretation of social meaning based on simple geometric figures.

**Results:** We found opposite neural responses in two core ToM regions : SCZ patients showed social content-related deactivation (relative to controls and BPD) of the right supramarginal gyrus, a region which activity is required to overcome egocentric “overmentalizing”, while the opposite pattern was found in BPD; reverse patterns, relative to controls and SCZ, were found in the left posterior superior temporal gyrus, a region involved in inferring other’s intentions. Receiver-operating-characteristic curve analysis showed 88% accuracy in discriminating the two clinical groups based on these neural responses.

**Conclusions:** These contrasting activation patterns of the temporoparietal junction in SCZ and BPD represent mechanistic differences of social cognitive dysfunction that may be explored as biomarkers or therapeutic targets.

## INTRODUCTION

An individual can represent abstractions of his relations with others and use them flexibly to guide interpersonal behaviors. This ability to recognize, handle and act accordingly with socially relevant information describes social cognition (SC)[1]. SC dysfunction, manifested by erroneous intention interpretations or idiosyncratic reactions to others’ emotions, has profound impact on functioning and daily life[2]. Neurocognitive processes of SC can be systematized in four main areas: emotion processing, social perception, attributional style and theory-of-mind (ToM)[3].

A dysfunction of ToM - the ability to infer others’ intentions, emotions and beliefs[4], has been described in severe mental disorders such as schizophrenia (SCZ), representing its most important predictor of functioning[5]. SC has also been implicated as a key contributor to functioning in bipolar disorder (BPD) patients [6, 7], and deficits were linked with psychosocial disadvantage [8]. Genetic studies have shown a complex polygenic picture for the architecture of psychiatric disorders [9]. Several common genetic variants were identified[10] that confer risk to both SCZ and BPD, as well as to autism spectrum disorders – the archetype of SC dysfunction – and current evidence favors a differentiation between SCZ and BPD that is more dimensional than categorical[11]. However, to our knowledge, no study has so far focused on the critical distinctions between neural processing of SC in these conditions.

The neural basis of SC involves a complex network of brain areas: ventromedial and dorsolateral prefrontal cortex (PFC), anterior cingulate cortex (ACC), amygdala, temporoparietal junction (TPJ) and insula[6, 12]. Meta-analyses of functional neuroimaging studies using ToM paradigms identified the TPJ and medial PFC as core areas, amongst other regions associated with the mirror neuron system (MNS)[13, 14]. Methodological considerations partly explain heterogeneous findings in ToM studies, but the putative existence of two ToM pathways is relevant: (1) implicit ToM, arising in early development (first 2 years of life), allowing spontaneous mental-state tracking and rapid extraction of others’ mental states; (2) later developing explicit ToM, with conscious analyses of external mental states, supported on additional executive functions[15, 16]. Of ToM’s core regions, only the TPJ has been shown to contribute in explicit and implicit processing, as early as in 7-month-old infants[16, 17]. This area, namely the right TPJ, has been proposed as a cluster of subregions: anterior TPJ, posterior TPJ and inferior parietal lobe regions[18, 19]. The TPJ and posterior superior temporal sulcus (pSTS) - the TPJ-pSTS complex, a phylogenetically recent hub, coordinate multiple brain networks when exploring dynamic social scenes typical of human experience[20].

ToM’s neural basis has been studied in SCZ patients. Despite literature inconsistencies, a meta-analysis of 21 fMRI trials with ToM paradigms in SCZ reported hypoactivity of its core network, with abnormal hyperactivation of attentional networks, suggesting vicarious mechanisms to ameliorate behavioral performance, which is frequently below par[21, 22]. For BPD, there is limited data on functional neuroimaging about ToM; a relative general hypoactivation of the MNS has been suggested[23-25]. However, direct comparison of SC’s specific neural functional correlates in both SCZ and BPD is currently missing. Moreover, a neurobiological distinction of social cognition processing in these conditions remains to be identified. We hypothesized that key structures at a relatively early processing level in the social brain, namely the TPJ, should present distinctive features during tasks that require low-level social judgment. To test this hypothesis, we investigated the neural substrates of ToM in patients with SCZ and BPD, using a ToM task requiring social interpretation of simple geometric figures.

## METHODS AND MATHERIALS

### Participants

Right-handed patients with BPD (n=20) and SCZ (n=20) matched for age (18-54), gender and education were recruited from the outpatient setting of a large tertiary hospital. Handedness was assessed through the Edinburgh Handedness Inventory[26]. All patients had an ICD-10 diagnosis of BPD or SCZ confirmed through direct interview by an experienced psychiatrist and medical records reviewing. Clinical stability as an inclusion criteria was operationalized through unchanged medication for at least 3 months, with a similar period of clinical stability: sustained euthymia (Brief Psychiatric Rating Scale’s - BPRS[27] mania and depression items ≤1) in BPD patients, and, in SCZ, Positive and Negative Symptoms Scale for Schizophrenia - PANSS[28] - score variation under 10%. Exclusion criteria included medical or neurological conditions (e.g. epilepsy, head trauma), comorbid alcohol or other drugs abuse/dependence, and MRI contraindications. The study was approved by the Ethics Commission of the Faculty of Medicine of the University of Coimbra (ref. CE-010/2014),conducted in accordance with the Declaration of Helsinki. All participants provided written informed consent to participate. Twenty right-handed healthy controls matched for age, gender and education were recruited from hospital and faculty workers and their relatives; only subjects with no personal or first-degree family history of psychiatric disorders were included.

### Neuropsychological assessments

Patients were assessed with the Insight and Treatment Attitudes Questionnaire (ITAQ)[29] and Personal and Social Performance Scale (PSP)[30], measuring, respectively, insight and social functioning. Patients and controls underwent neuropsychological assessment using the following SC psychometric instruments: Face Emotion Identification Test (FEIT)[31]; Social Perception Scale (SPS)[32]; Schema Component Sequencing Task – Revised (SCST)[33]; Ambiguous Intentions Hostility Questionnaire (AIHQ)[34]; Reading the Mind in the Eyes Test (RMET)[35]; Mentalization Questionnaire (MZQ)[36]; Toronto Empathy Questionnaire (TEQ)[37]. Evaluation was conducted within a 2-week interval after fMRI assessment.

### Stimuli Presentation and Apparatus

The experimental sessions were designed in Presentation software (version 17.0, Neurobehavioral Systems Inc., Albany, USA) and shown inside the MR scanner bore by means of an LCD screen (NNL LCD Monitor, NordicNeuroLab, Bergen, Norway; resolution 1920×1080, refresh rate 60 Hz) located approximately 156 cm awayParticipants viewed the screen through a mirror mounted above their eyes. Behavioral responses were collected using two fiber-optical MRI-compatible response pads, Lumina LS-PAIR (Cedrus Lumina LP-400, LU400 PAIR; Cedrus Corporation, San Pedro, USA), held in both hands.

### Experimental Design and Procedure

Participants were engaged in a visual task involving ToM animated stimuli[38, 39]. An experimental run consisted of 8 trials, each comprising a sequence of baseline, animation movie, jittering, and question blocks. The animation movie block, presented during 14s, displayed simple geometric shapes (two circles) on a bidimensional structured scenario. Prior to fMRI sessions, participants were told to interpret the animations as representing social interactions between two agents. There were four different animation categories: affiliative (friendly social interaction), antagonistic (hostile social interaction), indifferent (non-interacting social movements), linear (rectilinear non-social movements). Animations categories were presented twice per run, and animation blocks were preceded by a baseline block displayed for 12.5-13.5s, followed by a short jittering block (0-1s). Both blocks displayed a fixation circle. The variable duration of baseline and jittering blocks was used to ensure that fMRI volumes would not be acquired always in the same temporal instant of the different trials, so as to capture a more complete sampling of the hemodynamic response[40]. During question blocks an interrogative sentence was presented for 8.5s, asking participants to classify the emotional valence of social behaviors shown in preceding animations as positive (affiliative), negative (antagonistic), or indifferent (indifferent or linear animation). Participants were instructed to use the right index, right middle finger, and left index to indicate a positive, negative, or indifferent emotional valence, respectively. There were three different sets of randomized runs. Trios of participants (control, SCZ, BPD), matched during recruitment process, performed the same set of 4 randomized runs. Two SCZ participants performed two and three experimental runs, respectively, due to fatigue.

### fMRI Data acquisition and Pre-processing

Data were collected with a Siemens Magnetom TIM Trio 3T scanner (Siemens, Munich, Germany) with a phased array 12-channel birdcage head coil. The MR scanning session began by acquiring a 3-D anatomical T1-weighted MPRAGE (magnetization-prepared rapid gradient echo) pulse sequence (TR=2530ms; TE=3.42ms; TI=1100ms; flip angle 7°; 176 single-shot interleaved slices [no inter-slice gap] with voxel size 1×1×1mm; FOV 256mm). Functional images were acquired axially using a T2*-weighted gradient echo (GE) echo planar imaging (EPI) sequence covering the whole brain. Each functional series consisted of 150 volumes (TR=2000ms, TE=39ms, flip angle=90°, 29 interleaved slices [no inter-slice gap] with voxel size 3×3×4mm; FOV 254mm) of BOLD signal measurements.

### fMRI Data Analysis

Data processing was performed using BrainVoyager 21.2 (Brain Innovation, The Netherlands). Functional volumes pre-processing included slice-scanning time correction, interscan 3D head-motion correction, temporal high-pass filtering (GLM-Fourier, 2 cycles) and linear-trend removal. Functional data were normalized to Talairach anatomical space and spatially smoothed with a Gaussian kernel of 6-mm full-width at half-maximum.

For each experimental run, a general linear model (GLM) design matrix was defined, with box-car predictors for each experimental condition and confound predictors from head motion parameters and spikes. Predictors were convolved with the canonical hemodynamic response function (HRF). Then, resulting single-run beta values entered a second-level group random effects (RFX) GLM analysis.

### Statistical Data Analysis

Differences in age and education levels were assessed in patients (BPD or SCZ) and healthy participants through ANOVA. Independent samples t-test or chi-square comparisons of clinical data between BPD and SCZ patients were conducted.

Two behavioral measures of performance on the fMRI task were calculated: correct expected response percent score and average time spent to report a decision during the question block. For these variables, the effects of group and video animation category were assessed independently using Independent Samples Kruskal-Wallis H tests and Friedman tests, respectively.

Demographic, clinical and fMRI task behavior statistical analyses were performed using SPSS 24 (IBM, Armonk, USA).

To test our hypothesis of differential activation of the neural substrates underlying low-level ToM processing, a region of interest (ROI) was defined using a large bilateral TPJ mask in Talairach space. This extensive mask, with 25083 voxels, comprising a comprehensive set of ToM brain regions, was obtained by mirroring and converting to Talairach space[41] an existing right TPJ mask in MNI space[19]. A ROI voxel-wise RFX-GLM group analysis was conducted using functional data from all experimental runs. Resulting beta values were used to perform a RFX-ANOVA analysis with one within-subjects factor with repeated measures (category of animation video: affiliative, antagonistic, indifferent, linear) and one between-subjects factor (group: CTR, SCZ, BPD). RFX-ANOVA was used to test between-subjects’ differences, and the interaction between animation video category and group. Group level statistical F-maps were corrected for multiple comparisons using false discovery rate (FDR) method at q(FDR)<0.05.

In addition, a ROI RFX-GLM analysis was conducted within each cluster presenting a statistically significant interaction of category vs. group, to extract a single beta value for each predictor per subject. The receiver-operating-characteristic (ROC) analysis (sensitivity vs 1-specificity) was carried for each pairing of groups following univariate and multivariate classification approaches. The area under the ROC curve (AUC) was used as performance metric. The univariate approach, performed using SPSS 24, applied the ROC analysis independently to the beta values of the affiliative, antagonistic, indifferent, and linear predictors. The multivariate approach, implemented using MATLAB R2019 (MathWorks Inc., USA) and the Statistics and Machine Learning Toolbox, applied ROC analysis to the classification scores determined by binary linear support vector machine classifiers (10-fold cross validation, 50 runs) designed with four features corresponding to the beta values of the affiliative, antagonistic, indifferent, and linear predictors.

BOLD signal event-related average time courses were computed as follows: in each trial of the different animation video categories, percent BOLD signal change was determined using the start of the video as trigger, and as baseline the preceding period of 2 volumes (4 seconds of baseline block, displaying a fixation circle). Segments representing the same category of animation video were averaged over runs for the different groups of participants: controls, SCZ and BPD.

## RESULTS

Demographic and clinical data regarding participants can be found in **Table 1**.

**Table 1.** Demographic and psychopathologic sample characteristics.

| Table 1. Demographic and psychopathologic sample characteristics |  |  |  |  |  |
| --- | --- | --- | --- | --- | --- |
|  | Schizophrenia | Bipolar Disorder | Controls | Test statistics |  |
| Gender, male/female | 13/7 | 13/7 | 13/7 | N/A | N/A |
| Age, years (mean±SD) | 31.5±9.09; | 31.6±10.07 | 31.5±10,36 | F=0.001<sup>(a)</sup> | p=.999 |
| Education, years (mean±SD) | 13.6±3.68 | 13.85±2.64 | 14.9±4.52 | F=0.756<sup>(a)</sup> | p=.474 |
| Age of onset, years (mean± SD; range) | 26.06±7.05;<br>18-41 | 26.79±8.94;<br>16-50 | N/A | t=-0.276<sup>(b)</sup> | p=.784 |
| Duration of illness, years (mean±SD; range) | 4.33±5.20;<br>1-17 | 4.79±4.12;<br>1-16 | N/A | t=-0.297<sup>(b)</sup> | p=.769 |
| Number of lifetime admissions (mean±SD; range) | 1.00±1.49; 0-6 | 1.11±0.81;<br>0-3 | N/A | t=-0.268<sup>(b)</sup> | p=.790 |
| Antipsychotics, CPZE | 380.0<br>(50-1600) | 160.8<br>(0-1066) | N/A | t=2.226<sup>(b)</sup> | p=.032* |
| Psychotropic treatment, DDD (range) | 0.96<br>(50-1600) | 0.81<br>(0.00-2.00) | N/A | t=0.755<sup>(b)</sup> | p=.455 |
| History of psychosis, n (%) | 0.96 | 20 (100%) | N/A | χ<sup>2</sup>=0.035<sup>(c)</sup> | p=.106 |
| History of substance use, n (%) | (0.20-3.50) | 5 (25%) | N/A | χ<sup>2</sup>=0.557<sup>(c)</sup> | p=.731 |
| History of suicidal behaviors, n (%) | 16 (80%) | 4 (20%) | N/A | χ<sup>2</sup>=0.931<sup>(c)</sup> | p=1.000 |
| <sup>(a)</sup> ANOVA; <sup>(b)</sup> t Student test; <sup>(c)</sup> Chi square<br>BPD – bipolar disorder; CPZE - chlorpromazine equivalents; CTR – controls; DDD – defined daily dose;<br>SCZ – schizophrenia; SD – standard deviation |  |  |  |  |  |

### Psychopathology

Patients from both clinical groups were either in remission or sustained clinical stability, as shown by mean (SD) BPRS scores for patients with BPD [29.1(2.6)] and SCZ [35.6(6.4)]. Differences between two groups were statistically significant (p<.001). PANSS mean score in the SCZ sample was 44.7(SD=9.6). BPD patients had better social functioning compared to SCZ [PSP scores, 92.0(4.0)] vs 80.2(12.4), p=.001]. Insight was also lower in the SCZ sample: ITAQ mean of 17.1(3.2) vs. 19.1(2.2) in BPD patients.

### Neurocognition

In general, clinical groups had lower scores in SC measures, but differences were not significant compared to controls in facial emotion identification (FEIT), attributive style (AIHQ) and in the Reading-the-Mind-in-the-Eyes-Test (RMET).

The SCZ group had significantly lower performance than controls in social schemas assessment (SCST, p=0.015), empathy (TEQ, p=0.009), and emotional awareness (MZQ-EA subscale, p=0.002). Differences in social perception (SPS, p<0.001) between patients and controls (49.25±6.90) were particularly evident, with intermediate dysfunction found in BPD (39.90±6.09), and more severe impairment in SCZ (32.70±9.95).

Detailed neuropsychological results on SC can be found in **Table 2**.

**Table 2.** Social neurocognition evaluation results

| **Table 2. Social neurocognition evaluation results** |  |  |  |  |  |  |
| --- | --- | --- | --- | --- | --- | --- |
| TEST | Schizophrenia<br>(mean ± SD) | Bipolar<br>Disorder<br>(mean ± SD) | Controls<br>(mean ± SD) | ANOVA |  | Tukey Post Hoc<br>Test |
|  |  |  |  | F | p |  |
| FEIT | 11.95±2.96 | 12.30±2.77 | 13.75±2.38 | 2.467 | .094 |  |
| SPS | 32.70±9.95 | 39.90±6.09 | 49.25±6.90 | 22.482 | .000 | SCZ,BPD<CTR;<br>SCZ<CTR,BPD;<br>SCZ<BPD<CTR |
| SCST | 8.84±1.03 | 9.48±0.80 | 9.65±0.84 | 4.538 | .015 | SCZ<CTR |
| AIHQ | 2.53±0.76 | 2.27±0.75 | 2.47±0.63 | .689 | .506 |  |
| RMET | 23.10±3.32 | 21.90±4.74 | 24.55±4.19 | 2.068 | .136 |  |
| MZQ | 47.05±12.75 | 49.95±8.18 | 54.75±8.42 | 3.017 | .057 |  |
| MZQ-RS | 13.35±3.68 | 14.15±2.72 | 15.35±2.68 | 2.165 | .124 |  |
| MZQ-EA | 11.80±3.52 | 13.10±3.80 | 15.60±2.52 | 6.752 | .002 | SCZ<CTR |
| MZQ-PE | 11.65±3.60 | 12.95±3.28 | 13.10±3.16 | 1.130 | .330 |  |
| MZQ-RA | 10.25±3.16 | 9.75±2.51 | 10.70±2.23 | .638 | .532 |  |
| TEQ | 43.15±7.99 | 46.25±4.25 | 49.05±4.43 | . | .009 | SCZ<CTR |
| FEIT - Face Emotion Identification Test; SPS - Social Perception Scale; SCST - Schema Component Sequencing Task – Revised; AIHQ - Ambiguous Intentions Hostility Questionnaire; RMET - Reading the Mind in the Eyes Test; MZQ - Mentalization Questionnaire; MZQ subscales: RS – Refusing self-reflection, EA – Emotional awareness, PE – Psychic equivalence, RA – Regulation of affect; TEQ - Toronto Empathy Questionnaire; |  |  |  |  |  |  |

### ToM judgement behavior analysis

An Independent Samples Kruskal-Wallis H test revealed no statistically significant effect of group in expected response performance in controls (median=96.88%, IQR=5.65%) SCZ (median=94.79%, IQR=21.10%) and BPD (median=96.83%, IQR=14.84%); χ2(2)=3.783, p=0.151.

A statistically significant effect of animation category was found using a Friedman test, affiliative (median=100%, IQR=12.50%), antagonistic (median=100%, IQR=12.50%), indifferent (median=87.50%, IQR=25.00%), linear (median=100%, IQR=0%), χ2(3)=22.079, p<0.001. Post-hoc tests with Bonferroni correction for multiple comparisons indicated that participants had lower performance for indifferent than affiliative (p_Bonferroni_<0.05), antagonistic (p_Bonferroni_=0.05) or linear (p_Bonferroni_<0.05) animations.

Concerning time spent by participants to report the emotional valence of videos, an Independent Samples Kruskal-Wallis H test of behavioral data showed no significant effect of group (χ2(2)=1.309, p=0.520): controls (median=1176.19ms, IQR=559.16ms), SCZ (median=1286.22ms, IQR=256.87ms), BPD (median=1288.64ms, IQR=469.76ms). There was a statistically significant difference (χ2(3) = 26.980, p<0.001) in time spent to report decisions on different types of animation: affiliative (median=1057.57ms, IQR=526.09ms), antagonistic (median=1229.44ms, IQR=541.00ms), indifferent (median=1296.32ms, IQR=614.52ms), and linear (median=1225.00ms, IQR=533.50ms). Pairwise comparisons were performed with Bonferroni correction for multiple comparisons, showing that participants were quicker to report a decision after affiliative animations than indifferent (p_Bonferroni_<0.001) and linear blocks (p_Bonferroni_<0.05).

Illustrated ToM judgement behavioral data can be found in **Figure 1**.

**Figure 1.**
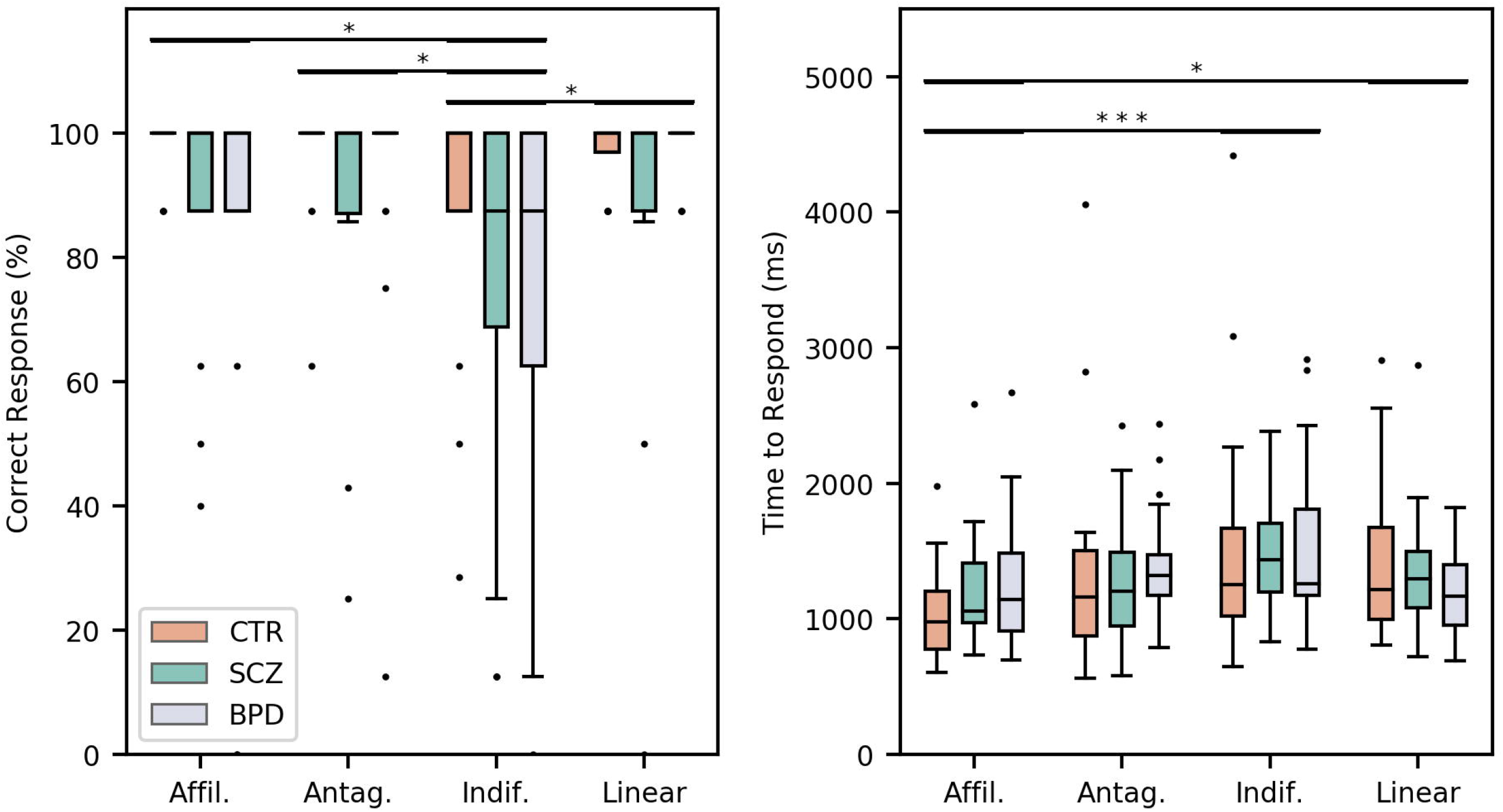
**Left** – correct response rate for different stimuli (affiliative, antagonistic, indifferent, linear). **Right** - mean time spent to report a decision in the decision block. CTR – controls; SCZ – schizophrenia; BPD – bipolar disorder.

### ToM judgment fMRI data

The ROI voxel-wise statistical group analysis testing the interaction between animation video category and group revealed a significant effect [F(6,171), q(FDR)<0.05, p<0.000531] in two groups of contiguous voxels, labelled as Clusters A and B. *Cluster A* comprises a region belonging to the right supramarginal gyrus (Brodmann area 40), whereas *Cluster B* includes part of the left posterior superior temporal gyrus (Brodmann area 22)[42]; these regions are presented in **Figure 2** and described in **Table 3**. Notably, several voxels of Cluster A and Cluster B revealed significant interaction effect after a stringent Bonferroni correction for multiple comparisons [F(6,171), p(Bonf)<0.05, p<0.000054]. Event-related time course analyses of the two clusters are shown in **Figure 2**, showing distinct mean percent BOLD signal change for controls, SCZ and BPD, during affiliative, antagonistic, indifferent and linear animations. Concerning main group effects, ROI voxel-wise statistical analysis didn’t show significant differences [F(2,57),q(FDR)>0.05, p>0.001030] between controls, SCZ and BPD.

**Table 3.** Summary of the ROI voxel-wise statistical group analysis testing the interaction between the category of animation video and group (q(FDR)<0.05, P<0.000531)

|  |  |  |  |  |  |  |
| --- | --- | --- | --- | --- | --- | --- |
| Region | No. of Voxels* | Peak voxel coordinates<br>(Talairach) |  |  | F(6,171) | P-value |
|  |  | X | Y | Z |  |  |
| Cluster A | 74 (6) | 63 | -46 | 25 | 5.564 | 0.000027 |
| Cluster B | 82 (4) | -66 | -43 | 16 | 5.286 | 0.000050 |
| *The number of voxels is based on the resolution of the anatomical dataset 1x1x1mm<sup>3</sup>.The values in parentheses are the number of significant voxels after Bonferroni correction for multiple comparisons, p(Bonf)<0.05. |  |  |  |  |  |  |

**Figure 2.**
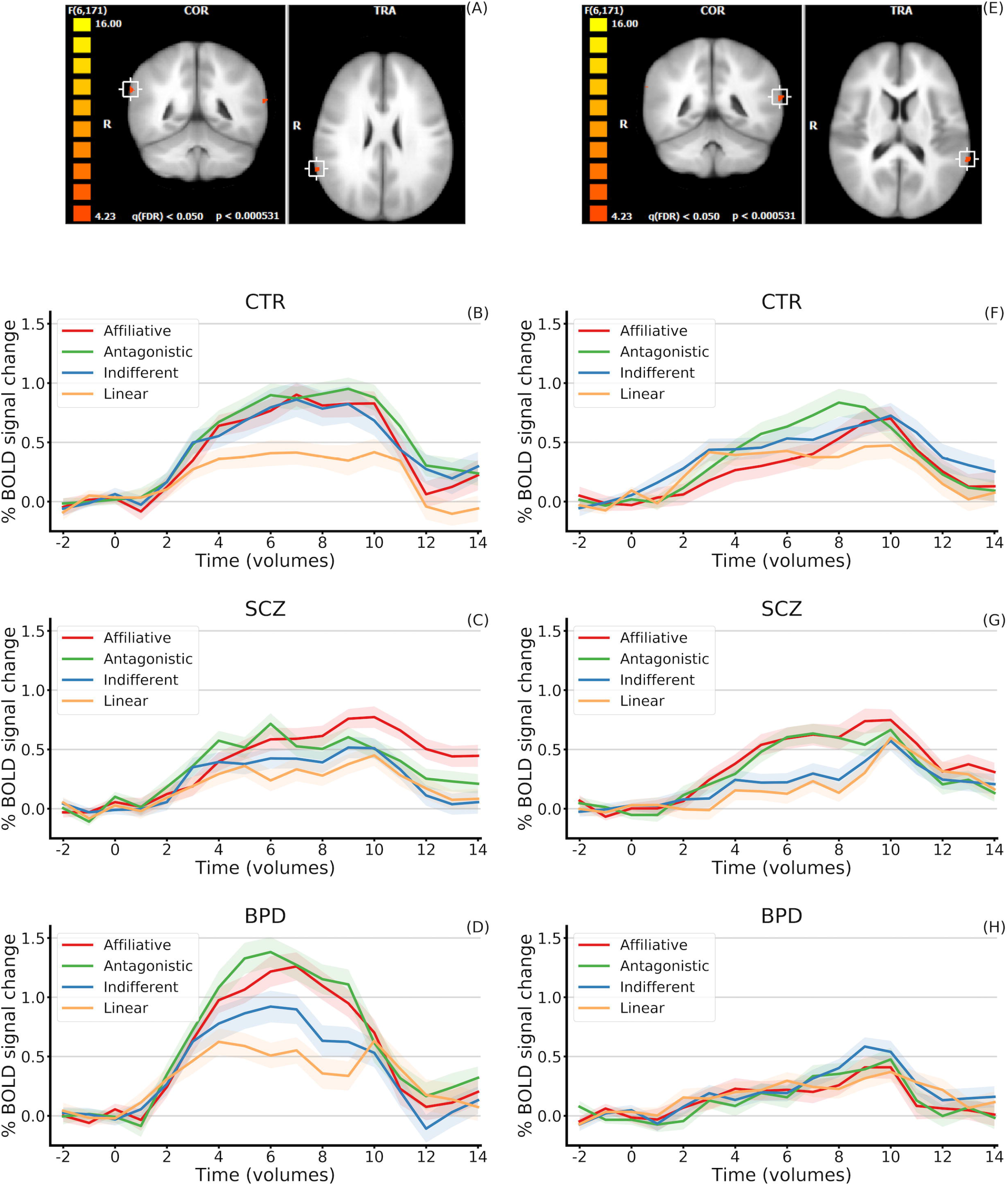
**A and E**. Result of RFX-ANOVA analysis. Interaction Group x Stimuli. A – right supramarginal gyrus (Brodmann area 40); B – left posterior superior temporal gyrus (Brodmann area 22). **B, C and D**. Event-related mean percent BOLD signal change time courses in Cluster A, right supramarginal gyrus, BA40, for the different stimuli. Shaded regions represent the standard error. **F, G and H**. Event-related mean percent BOLD signal change time courses in Cluster B, left posterior superior temporal gyrus, BA22, for the different stimuli. Shaded regions represent the standard error. CTR – controls; SCZ – schizophrenia; BPD – bipolar disorder.

A ROC analysis showed that single univariate classification by activation in the right supramarginal gyrus provided the best discrimination. Detailed results can be found in **Table 4**, with ROC analysis, and **Figure 3**, with Beta values of the affiliative, antagonistic, indifferent, and linear predictors in Clusters A (right supramarginal gyrus) and B (left posterior superior temporal gyrus).

**Table 4.** Classification performance expressed using area under the receiver operating characteristics curve

|  |  | Cluster A<br>Right Supramarginal Gyrus |  |  | Cluster B<br>Left posterior Superior Temporal Gyrus |  |  |
| --- | --- | --- | --- | --- | --- | --- | --- |
|  |  | CTR vs SCZ | CTR vs BPD | SCZ vs BPD | CTR vs SCZ | CTR vs BPD | SCZ vs BPD |
| Univariate | *Affil.* | 0.713<br>(0.090) | 0.633<br>(0.089) | 0.880<br>(0.054) | 0.570<br>(0.093) | 0.540<br>(0.093) | 0.650<br>(0.091) |
|  | *Antag.* | 0.620<br>(0.093) | 0.643<br>(0.087) | 0.788<br>(0.072) | 0.513<br>(0.093) | 0.620<br>(0.095) | 0.628<br>(0.092) |
|  | *Indif.* | 0.570<br>(0.094) | 0.598<br>(0.091) | 0.703<br>(0.087) | 0.595<br>(0.092) | 0.608<br>(0.094) | 0.545<br>(0.094) |
|  | *Linear* | 0.570<br>(0.092) | 0.603<br>(0.092) | 0.683<br>(0.088) | 0.645<br>(0.088) | 0.540<br>(0.094) | 0.603<br>(0.092) |
| Multivariate |  | 0.651<br>(0.003) | 0.567<br>(0.007) | 0.858<br>(0.001) | 0.776<br>(0.002) | 0.599<br>(0.004) | 0.694<br>(0.003) |

**Figure 3.**
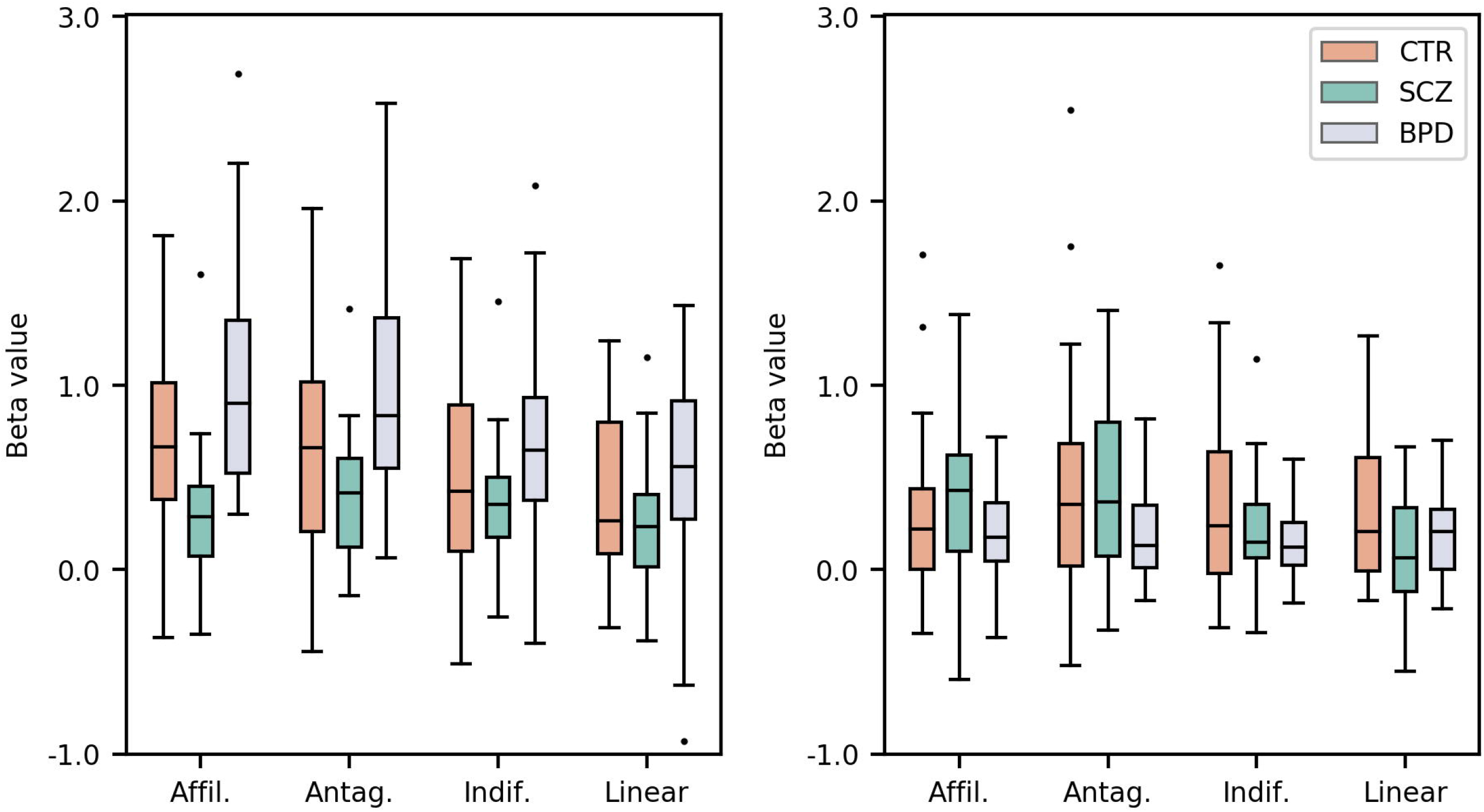
Beta values of the affiliative, antagonistic, indifferent, and linear predictors in the Cluster A - right supramarginal gyrus (left) and Cluster B - left posterior superior temporal gyrus (right), for the CTR, SCZ, and BPD groups.

Interestingly, performance was best at discriminating between clinical groups, reaching an AUC of 0.88 for positive stimuli and 0.79 for negative. The ROC analysis of the multivariate classification scores also showed a high AUC at discriminating SCZ and BPD (0.86) using features from the right supramarginal gyrus cluster. The multivariate approach designed with the left posterior superior temporal gyrus features reached an AUC of 0.78.

## DISCUSSION

The main novelty of this study lies in the discovery of a fundamental neurobiological difference in early social cognition processing between SCZ and BD, using a simple social animation paradigm.

We found a critical functional dissociation in low-level ToM in BPD vs. SCZ, with contrasting activation patterns in two core regions with the TPJ complex: the right supramarginal gyrus (SMG) and left posterior superior temporal gyrus (STG). This startling finding was observed in patient groups of short disease duration, with slightly subpar performance in some behavioral and neuropsychological SC measures compared to controls.

Activation of the TPJ, but not of other relevant ToM regions (e.g. PFC), has been found with near-infrared spectroscopy in 7-month-old babies, using ToM visual paradigms, suggesting infants already draw on this region for implicit ToM, similarly to what adults do when reasoning explicitly[17]. Our paradigm, focused on low-level perceptual evaluation, was expected to elicit implicit ToM pathways. These emerge relatively early in both phylogenetic grounds and human neurodevelopment; the right SMG has been known to activate more significantly in somatoperceptive ToM stories, while the more posterior TPJ is associated with ideational story stimuli[43].

The nature of differences driving the interaction between group and low-level video categories could relate to right SMG’s (which we found to have reduced activity in SCZ and hyperactivity in BPD) role in overcoming the egocentric bias (“overmentalizing”)[44]. Conversely, the left posterior STG (underactivated in BPD) is involved in inferring other’s intentions and introspective accuracy[45]. These findings suggest contrasting SC neural dysfunctions in these disorders, as confirmed by our ROC analysis, with robust discriminative power of these regions’ activation in distinguishing BPD and SCZ, especially for positive stimuli in the right SMG.

In psychosis, as in autism, there is difficulty setting aside self-knowledge to appreciate another person’s subjective world, and it has been so far believed that psychotic and mood conditions were diametric to autism spectrum disorders, associated, respectively, with hyper- or hypofunctioning aspects of SC[46].

Our results bring a new perspective to this view, by showing a critical distinction between SCZ and BPD. This suggests that addressing mentalization as a unidimensional continuum of intercorrelated functions across these disorders could be methodologically fallacious[47]. ToM dysfunction in SCZ patients could reflect impaired perspective-taking, explaining the egocentric projection of patients’ suspicions and biases onto others in ambiguous situations - “over-mentalizing”, yet failing to grasp the interplay between different perspectives in social interactions - “under-mentalizing”[47]. Our study suggests a more fundamental and distinct deficit across conditions occurring even in simple social animations.

Present findings of a dissociation between neural responses of SCZ and BPD in the right SMG - Brodmann area 40 (BA40) - underpin the relevance of this area to ToM processing. An elegant multimodal study, using fMRI and transcranial magnetic stimulation, found that disruption of right BA40 favored one’s emotional egocentricity bias –tendency to use the self as a reference point when perceiving the world and emulating others’ mental states; right SMG normal activation would be valuable to avoid biased social judgements[44]. Independent structural and functional connectivity studies have supported parcellation of the right TPJ, with anterior TPJ, which includes the right SMG, exhibiting robust connectivity with areas relevant to socioemotional processing (e.g. ACC,insula); on the other hand, TPJ posterior areas, including the angular gyrus, have stronger connectivity with the medial PFC[18, 19, 44].

The differential activation of right SMG between BPD and SCZ hereby found is consistent with previous findings regarding degree centrality (DC) - studying the number of instantaneous functional connections between a region and the rest of the brain[48]: in SCZ and psychotic BPD patients, using DC from an executive/working-memory fMRI task, DC increase in BPD, compared with SCZ, was found in the right SMG.

The right lateralization of SMG findings is anatomically supported by current knowledge regarding von Economo’s neurons (VEN), key functional units to SC and the default-mode network. A study exploring functional connectivity in resting-state of VEN-rich areas found significantly asymmetric preponderance of right vs. left SMG (74 vs. 26%), something that was not found in other areas, other than the functionally associated right TPJ complex[49].

In SCZ, where a relative inversion of usual left lateralization of temporal lobe response to language has been reported, it was hypothesized that emotional prosody response could also been inverted, lateralized to the left hemisphere[50, 51]; such a double inversion was found in a case report after epilepsy neurosurgery[52]. Early neurodevelopmental changes could determine interhemispheric compensation mechanisms. A fMRI study in SCZ and BPD reported left lateralization of response to emotional prosody, along with left SMG hypoactivation in BPD compared to SCZ, and right SMG hypoactivation in BPD compared with controls[51]. Stimuli, unlike our study, were auditory: participants listened to sentences describing happy or sad scenarios.

Morphometric studies pinpointed the SMG as relevant to SCZ and BPD’s neuropathology, with volumetric reduction[53-55], predominantly in SCZ. Diffusion-tensor imaging also found SMG changes: reduced and augmented fractional anisotropy in SCZ and BPD, respectively, with increased mean diffusivity of grey matter in both[56]. In a recent study with innovative morphometric measures such as gyrification, exploring the present sample, we found increased SMG gyrification in BPD, while in SCZ it was reduced[57].

A second cluster of differential activation was found in the left posterior STG, with reverse patterns: increased and attenuated response in SCZ and BPD, respectively. This region, home to Wernicke’s area, corresponds to the posterior section of Brodmann area 22 (BA22), ventral to the SMG. The superior temporal sulcus (STS) is implicated in understanding others’ intentionality, and inferential processing (ToM) has been associated with the BA22[58].

A meta-analysis of functional imaging studies of emotional perception in SCZ reported greater activation in the posterior STG[59]. However, reduced activation of the left STG/BA22 during SC paradigms has been found during acute SCZ episodes[60]. A recent fMRI study investigated the neural correlates of SC introspective accuracy (IA) in SCZ, measuring activity during a facial emotion recognition task requiring contemporaneous self-assessments; SCZ patients showed reductions in IA-specific neural activity of critical regions for IA, when compared with controls [45]. Analysis comparing activation in SCZ of IA-specific neural activity vs. a control task reported the highest activation difference in the right STG/BA22 (peak Z=3.95). Additionally, the left BA22 (peak Z=3.34) was one of the regions with stronger positive correlation between IA ability and neural activation, in healthy individuals, relative to SCZ.

In a recent meta-analysis of whole-brain voxel-based morphometry studies in BPD, right STG volume was smaller in some subgroups: adult and type I BPD patients[61]. An fMRI study found increased activation of left STS in BPD compared to controls and SCZ; this, along with other activation data during emotion processing in BPD, has been interpreted as an excessively activated emotion-processing system, with heightened significance attributed to emotional stimuli, generating hypervalent affective states[62, 63].

In general, ‘hyper-empathy’ in BPD, with a trend towards better ‘affective’ than ‘cognitive’ empathy, might equate with known PFC dysfunction and relatively preserved limbic function in BPD[23, 64]. Non-surprisingly, ToM impairments have been suggested as a BPD trait marker, related neither with illness years nor medication, or state-related variables such acute phases[25, 65].

Despite our case-control study’s strict design, namely inclusion/exclusion criteria and matching relevant variables (age, gender, education), with comparable clinical characteristics (e.g. disease years, substance abuse), some limitations should be discussed. First, its cross-sectional nature. Also, sample size precludes some assessments, namely the influence of psychotic symptoms in BPD. While we assessed current medication, its possible effect as a confounder (e.g. antipsychotics exposure) cannot be excluded. Nonetheless, opposite neural changes documented in BPD and SCZ patients could not be attributed to current antipsychotic usage, which was intermediate in BPD, high in SCZ, and absent in controls.

Overall, in this functional neuroimaging study we found a neural response dissociation in BPD vs. SCZ during low-level processing in core ToM regions, with contrasting activation patterns found in right SMG and left posterior STG. This shows a fundamental distinction in social cognition processing ocurring at a relatively early level in these conditions and that had so far remained unsuspected. This dissociation, between well-matched patients of two disorders sharing morphometric and genetic features[66], sheds light to characteristic neural basis of impaired social cognition in SCZ and BPD, where such deficits could be the most significant predictor of functionality[5, 8]. While a perceived feature of psychiatric neuroimaging research has been its relative disconnection from healthcare and its unmet needs[67], these findings could translate into meaningful gains to clinical practice, given the promising ROC analysis results. This novel feature of opposing TPJ activation patterns could be investigated as a potential biomarker, or even as an anatomical target for differential therapeutic neuromodulation, besides highlighting this neural structure’s relevance to the social challenges faced by these patients in daily life.

## Data Availability

The datasets generated during and/or analysed during the current study are available from the corresponding author on reasonable request.

## ACKNOWLEDGMENTS

Financial support for the conduct of this research was granted by the Portuguese Foundation for Science and Technology (FCT), through the projects ‘From molecules to man: novel diagnostic imaging tools in neurological and psychiatric disorders’ (reference CENTRO-07-ST24-FEDER-00205 and BIGDATIMAGE, CENTRO-01-0145-FEDER-000016), ‘MEDPERSYST: Synaptic networks and Personalized Medicine Approaches to Understand Neurobehavioral Diseases Across the Lifespan’ (reference SAICTPAC/0010/2015, POCI-01-0145-FEDER-016428), PTDC/PSI-GER/1326/2020 and FCT UID/4950/2020, FCT also funded individual grants to JVD (Individual Scientific Employment Stimulus 2017 - CEECIND/00581/2017) and RM (Institutional Call to Scientific Employment Stimulus 2018 - CEECINST/00041/2018).

We would like to thank the participants for their involvement in this study. We are also very grateful to: Paula Tavares for the ToM visual animation task; Carlos Ferreira and Sónia Afonso for the help with MRI setup and scanning; Licínio Craveiro for English language reviewing.

## DISCLOSURES

The authors declare no financial disclosures or competing interests. A preliminary version of this manuscript has been posted in a preprint server – medRxiv.

## Notes

### Competing Interest Statement

The authors have declared no competing interest.

### Author Declarations

The study was approved by the Ethics Commission of the Faculty of Medicine of the University of Coimbra (ref. CE-010/2014), conducted in accordance with the Declaration of Helsinki.

